# Deciphering the tangible spatio-temporal spread of a 25 years tuberculosis outbreak boosted by social determinants

**DOI:** 10.1101/2022.02.07.22270243

**Authors:** Mariana G. López, Mª Isolina Campos-Herrero, Manuela Torres-Puente, Fernando Cañas, Jessica Comín, Rodolfo Copado, Penelope Wintringer, Zamin Iqbal, Eduardo Lagarejos, Miguel Moreno-Molina, Laura Pérez-Lago, Berta Pino, Laura Sante, Darío García de Viedma, Sofía Samper, Iñaki Comas

## Abstract

**Background:** Outbreak strains are good candidates to look for intrinsic transmissibility as they are responsible for a large number of cases with sustained transmission. However, assessment of the success of long-lived outbreak strains has been flawed by the use of low-resolution typing methods and restricted geographical investigations. We now have the potential to address the nature of outbreak strains by combining large genomic datasets and phylodynamic approaches.

**Methods:** We retrospectively sequenced the whole genome of representative samples assigned to an outbreak circulating in the Canary Islands (GC) since 1993; accounting for ∼20% of local TB cases. We selected a panel of specific SNP markers to in-silico search for additional outbreak related sequences within publicly-available TB genomic data. Using this information we inferred the origin, spread and epidemiological parameters of the GC-outbreak.

**Findings:** Our approach allowed us to accurately trace both the historical and recent dispersion of the strain. We evidenced its high success within the Canarian archipelago but found a limited expansion abroad. Estimation of epidemiological parameters from genomic data contradicts a distinct biology of the GC-strain.

**Interpretation:** With the increasing availability of genomic data allowing for an accurate inference of strain spread and key epidemiological parameters, we can now revisit the link between *Mycobacterium tuberculosis* genotypes and transmission, as routinely done for SARS-CoV-2 variants of concern. We show that the success of the GC-strain is better explained by social determinants rather than intrinsically higher bacterial transmissibility. Our approach can be used to trace and characterize strains of interest worldwide.

**Funding:** European Research Council (101001038-TB-RECONNECT), the Ministerio de Economía, Industria y Competitividad (PID2019-104477RB-I00), Instituto de Salud Carlos III (FIS18/0336), European Commission –NextGenerationEU (Regulation EU 2020/2094), through CSIC’s Global Health Platform (PTI Salud Global) to IC. Gobierno de Aragón/Fondo Social Europeo “Construyendo Europa desde Aragón” to SS

**Research in context:** *Evidence before this study:* Identification of intrinsically highly transmissible strains of *Mycobacterium tuberculosis* remains elusive. Among candidates are those strains that have been thriving in a community for decades representing a significant contribution to the long-term local TB burden. These long-lived outbreak strains have been identified in different parts of the world and the speculation is that their success is linked to higher transmissibility. Several studies have attempted to analyze the epidemiological characteristics of these strains as well as their genomic composition to look for potential transmission determinants. However those studies are usually circunscribe to their original geographic boundaries. By contrast, this transmissibility should be replicated in different parts of the world, a lesson learnt from SARS-CoV-2 variants of concern. Previous attempts failed to examine the success of these outbreak strains at a global scale. Thus, it is unknown whether the long-lived outbreak strains had a similar or different trajectory in other countries, casting doubts about their transmissibility potential.

*Added value of this study:* Here we analyzed a strain causing a long-lived outbreak in the Canary Islands since 1993 using whole genome sequencing. As in previous studies with other similar outbreak strains, we analyzed the diversity and phylodynamics of the outbreak in the area where it was originally described. However, thanks to the possibility of interrogating the entire European Nucleotide Archive, we had the unique chance to look at the spread of the strains beyond its original geographic boundaries. This approach allowed us to comprehensively trace the real spatio-temporal spread of the outbreak from the emergence of its ancestor about 700 years ago to its recent transmission outside the Canary Islands. As a result, there is limited evidence for similar success of the strains outside Canary Islands. Furthermore, we complemented the analysis with epidemiological data of the early cases and with phylodynamic analysis to estimate key epidemiological parameters linked to the strain spread. All evidence strongly suggests that factors related to the host, instead of the bacteria, are behind the persistence and expansion of the outbreak strain.

*Implications of all the available evidence:* Infectious disease outbreaks are a major problem for public health. Tracing outbreak expansion and knowing the main factors behind their emergence and persistence are key to an effective disease control. Our study allows researchers and public health authorities to use WGS-based methods to trace outbreaks, and include available epidemiological information to evaluate the factors underpinning outbreak persistence. Taking advantage of all the information freely available in public repositories, researchers can accurately establish the expansion of the outbreak behind its original boundaries; and they can determine the potential risk of the strain to inform health authorities which, in turn, can define target strategies to mitigate its expansion and persistence. Finally, we show the need to evaluate strain transmissibility in different geographic contexts to unequivocally associate its spread to local or pathogen factors, a major lesson taken from SARS-CoV-2 genomic surveillance.

## Introduction

Tuberculosis (TB) has been the first cause of death by an infectious disease for years surpassing HIV according to the World Health Organization (WHO). In 2019 were reported 10 million new TB cases and 1.4 million deaths [1], with these numbers likely to increase due to the COVID-19 pandemic [2]. Outbreaks, defined as the concentration of an abnormal number of disease cases in space and time, are very common in TB. While outbreaks have generally been assumed to be short-lived, genotyping has been able to identify outbreak strains that became highly prevalent in specific regions and end up transmitting over decades. These outbreak strains can be found worldwide and across MTBC lineages and are usually drivers of local TB burden [3–8]. However, success of outbreak strains has been largely evaluated only in their original location, raising the question of whether those strains have any intrinsic transmissibility advantage or if their success is associated with local population processes like founder effects or ecological drivers of transmission [3–8].

In this work we investigate in detail one of those outbreak strains and compare them to others, previously published, to learn about the origin and epidemiology of long-lived strains. In 2001 a TB outbreak was identified in the Canary Islands. 651 strains were retrospectively analyzed between 1991-1996 in Gran Canaria (GC), the most populated island of the archipelago, using RFLP IS*6110*. A big cluster of 75 isolates was recognized with the first 10 cases diagnosed in 1993. The likely index case was a Liberian refugee who arrived on the island 6 months before diagnosis, in July of 1993 [9]. The strain belongs to the Beijing genotype, corroborated by spoligotyping, and was named GC1237. More recently, three GC-outbreak single nucleotide polymorphisms (SNPs) were selected and used to design a specific PCR to rapidly identify secondary cases belonging to GC-outbreak [10]. The fast spread of the GC strain to the other islands of the Canarian archipelago was evidenced by using different molecular typing methods, and also new cases outside the archipelago were identified up to 2014 [10–12]. Until now, GC-outbreak has never been studied by exploring the information obtained from whole genome sequencing (WGS).

In this work, we track the outbreak strain in its original geographic boundaries but also elsewhere by querying the entire European Nucleotide Archive (ENA) database. Then, we combine epidemiological and sequencing data, and apply a phylodynamic analysis to trace the origin of the GC outbreak strain, to track its spread, to understand its dynamics and to define the factors that underpin its expansion.

## Materials and method

### Study population

*M. tuberculosis* DNA samples from 80 different patients, 3 additional ones from the index case, all from Canary Islands, and 1 from Zaragoza were supplied by the Instituto de Investigación Sanitaria de Aragón. In addition, DNA samples from 5 different patients from the peninsula (Madrid) were supplied by the Hospital Universitario Gregorio Marañón, 2 of which belonged to recent immigrants from Guinea (Table S1). A representative number of samples from the Canary Islands, ranging the defined period of the outbreak (1993-2014), was selected. All samples were previously identified as part of the Gran Canaria (GC) outbreak by genotyping methods.

Epidemiological information was obtained from Servicio de Microbiología (Hospital Universitario de Gran Canaria Dr. Negrín) under the Ethics Committee Application CEIm H.U.G.C. Dr. Negrín 2019-502-1.

### Study design

The whole genome of *M. tuberculosis* from the first set of 89 samples (86 patients) was sequenced. Additional samples were looked for by querying our local database of sequences [13–15], with the three GC outbreak specific SNPs [10] previously selected; Rv2524 (C1398T, SNP2847935), Rv3869 (G1347C, SNP4346385) and Rv0926c (G162A, SNP1033625). Redefinition of specific SNPs was conducted based on phylogeny and distance analysis. New samples were identified by inspecting the whole ENA repository and samples from ongoing projects. Phylogeographic and phylodynamics analysis were performed in order to evaluate the outbreak. Comparison with other outbreaks was performed; sequences corresponding to Denmark (PRJEB20214)[4]; Thailand (PRJN244659)[3]; Bern (PRJEB5925)[6]; and Buenos Aires (BA, PRJEB7669)[5] outbreaks were downloaded from ENA and analyzed with the bioinformatics pipeline detailed below.

### Whole-Genome Sequencing and bioinformatics analysis

DNA samples were used to prepare sequencing libraries with a Nextera XT DNA library preparation kit (Illumina), following the manufacturer’s instructions. Sequencing was performed on an Illumina MiSeq instrument, applying a 2 × 300bp paired-end chemistry. General bioinformatics analysis is described in https://gitlab.com/tbgenomicsunit/ThePipeline/-/tree/master/. Briefly, read files were trimmed and filtered with fastp [16]. Kraken software was used to remove nonMTBC (non-Mycobacterium tuberculosis complex) reads [17]. Sequences were then mapped to an inferred MTBC common ancestor genome (https://doi.org/10.5281/zenodo.3497110) using bwa [18]. SNPs were called with SAMtools [19] and VarScan2 [20]. GATK HaplotypeCaller [21] was used for InDels calling. SNPs with a minimum of 20 reads (20X) in both strands and quality 20 were selected. InDels with less than 20X were discarded. SnpEff was used for SNP annotation using the H37Rv annotation reference (AL123456.2). Finally, SNPs falling in genes annotated as PE/PPE/PGRS, ‘maturase’, ‘phage’, ‘13E12 repeat family protein’; those located in insertion sequences; those within InDels or in higher density regions (>3 SNPs in 10 bp) were removed due to the uncertainty of mapping. Heterogenous SNPs (hSNPs) were obtained from filtered SNP files, they were classified as positions were > 5% and < 90% of the reads were the alternative allele [22]; we looked for hSNPs only for positions for which at least one sample harbor the SNP fixed, i.e were > 90% of reads were the alternative allele. Lineages were determined by comparing called SNPs with specific phylogenetic positions established [23,24]. An in-house R script was used to detect mixed infections based on the frequency of lineage- and sublineage-specific positions [15]. Pairwise distances were computed with the R *ape* package, statistical analysis and graphics were also performed with R. Raw sequencing data are available under the accession number PRJEB50491 (ENA).

All WGS Illumina sequencing runs stored in the ENA with metadata identifying them as *M. tuberculosis* complex as of July 2018 were downloaded (N=38075). A de Bruijn graph (k=31) was built from each with Cortex v1.0.5.21 (https://github.com/iqbal-lab/cortex/releases/tag/v1.0.5.21), and sequencing errors were removed by excluding low coverage unitigs from the graph (threshold is sample dependent and automatically chosen by the software). The remaining kmers from these graphs were then used to build a BItsliced Genomic Signature Index (BIGSI)[25]. The index was used to query these ENA samples for the outbreak-related SNPs, by first creating two 61 base-pair probes for each SNP (30bp flanking each side of the SNP, from the reference genome, and one probe for each SNP allele), returning binary information as to which sequencing runs contained all the kmers in the probes.

### Phylogenetic analysis

Multisequence alignment (MSA) files were constructed with concatenated SNPs discarding well-known drug resistance and invariant positions. Maximum likelihood trees were inferred with RAxML v8.2.11 [26] using GTRCATI model and 1000 fast-bootstrap replicates. Tree visualization and editing were conducted in ITOL (https://itol.embl.de/). Specific SNPs were identified using likelihood ancestral reconstruction of Mesquite software v3.61 (http://www.mesquiteproject.org). Genomic network was constructed with the MSA files with a median joining network inference method implemented in PopArt Software [27].

### Time and geographic origin of the outbreak

A set of 200 samples including the outbreak, closest clades and a representative subset of global samples selected with Treemer [28] was used to estimate the time of the most common ancestor (tMRCA) of the outbreak and deeper nodes. The evolutionary history of the outbreak was reconstructed with BEAST2 v2.5.1 [29] using GTR (gamma 4) as site model, coalescent constant population as tree prior and relaxed clock log-normal with gamma distribution as prior. We use a tip dating method and also set the clock rate (mean = 4.6 × 10^−8^ substitutions per site per year with 95% HPD: 3.3 × 10^−8^ to 6.2 × 10^−8^), based on previous publications [30]. Ascertainment bias was corrected by adjusting the clock rate based on the size of the alignment [14]. MCMC’s chain length of 10M with sampling every 1000 steps for the posterior distribution. Three independent runs were performed to reach convergence. Log and tree files were combined with LogCombiner tool discarding 10% of burn-in. Combined files were inspected with Tracer v1.7.1 [31], all parameters reached effective sample size (ESS) > 200 and well mixing. Tree was annotated with TreeAnnotator and visualized with FigTree v1.4.3.

Spatial dispersal dynamics of the outbreak and closest clades was reconstructed using a phylogeographic diffusion in discrete space approach implemented in BEAST v2.5.1 (Beast-classic package) [32]. A set of 142 samples, including the whole outbreak and closest clades, and those global strains with available information of year and location were used. Same clock and tree model, as well as priors, as in dating analysis were used. Same MCMC chains and further analysis as dating were conducted. In addition, SPREAD3 tool was used for geographic visualization of outbreak evolutionary history [33], only dispersion patterns with posterior values higher than 0.9 were plotted.

### Phylodynamics of the outbreak

The dynamics of the outbreak was studied with the Bayesian Birth-Death Skyline model implemented in BEAST2 v2.5.1. This model allows estimating parameters related to the evolution of the outbreak [34]. The analysis was conducted with all the samples included in the outbreak (64 samples, Table S1). We use a GTR (gamma 4) as site model, a strict6000 clock with LogNormal prior with the mean previously published [30] as prior. A Birth death skyline serial tree model was used. Specific parameters were set as follow (i) *becoming uninfectious* (LogNormal; M=0, S=1.25), considering the global estimation of 1 year as TB infectiousness period and not taking into account latency [35] (ii) *origin* (LogNormal; M= 30; S=0.04) it was taking into account that the likely index case has arrived to the archipelago in the beginning of 1993 starting immediately the outbreak; (ii) *reproductive number* (LogNormal; M=0; S= 1.25; dimension = 13) it was considered that in settings with low TB burden each person can generate a maximum of 12 secondary cases [35]; (iv) *sampling proportion* (beta; alpha = 3; beta = 50) it was calculated as the total number of TB cases in Canary Islands between between 1993-2017 (https://www3.gobiernodecanarias.org/sanidad/scs/listaImagenes.jsp?idDocument=fe9e3e94-feee-11e0-ab85-376c664a882a&idCarpeta=0f67aaf7-9d88-11e0-b0dc-e55e53ccc42c) and considering that 20-30% of cases belong to the outbreak resulting in a sampling range of 8-13%. MCMC’s chain length of 10M was run with sampling every 1000 steps for the posterior distribution. Log file was inspected with Tracer v1.7.1 [31], all parameters reached ESS>200 and well mixing. Results were inspected and plotted with R package bdskytools (https://github.com/laduplessis/bdskytools).

Clocklike structure of the outbreak dataset was first evaluated with a linear regression analysis of tip dates vs root-to-tip distances with TempEst [36] and by applying a date randomization test (DRT) with 100 randomized replicates of the BDSKY. The clocklike structure was evaluated with the analysis proposed by [37] considering that DRT is passed when the clock rate estimate for the observed data does not overlap with the range of estimates obtained from the randomized sets; intermediate DRT is passed when the clock rate estimate for the observed data does not overlap with the confidence intervals of the estimates obtained from the randomized sets, and stringent DRT is passed when the confidence interval of the clock rate estimate for the observed data does not overlap with the confidence intervals of the estimates obtained from the randomized sets.

### Data and code availability

All sequences have been deposited on the ENA repository under the project number PRJEB50491. All the scripts, tools and reference sequences are included in the repository section of the laboratory webpage (http://tgu.ibv.csic.es/?page_id=1794) and the gitlab page (https://gitlab.com/tbgenomicsunit/ThePipeline). Project numbers of the additional sequences from other studies are listed in Tables S2 and S4. Any additional information required to reanalyze the data reported in this paper is available from the lead contact upon request.

## Results

### Genomic delineation of the GC outbreak

We retrieved 86 samples from outbreak patients previously typified by molecular methods, plus 3 additional samples from the likely index case collected in different years. Out of those, we discarded 24 samples which did not achieve sufficient DNA quality for sequencing, leaving us with a total of 65 usable samples (62 patients, ∼10% of the outbreak, Table S1). The GC outbreak was delineated following the diagram detailed in Figure 1.

**Figure 1.**
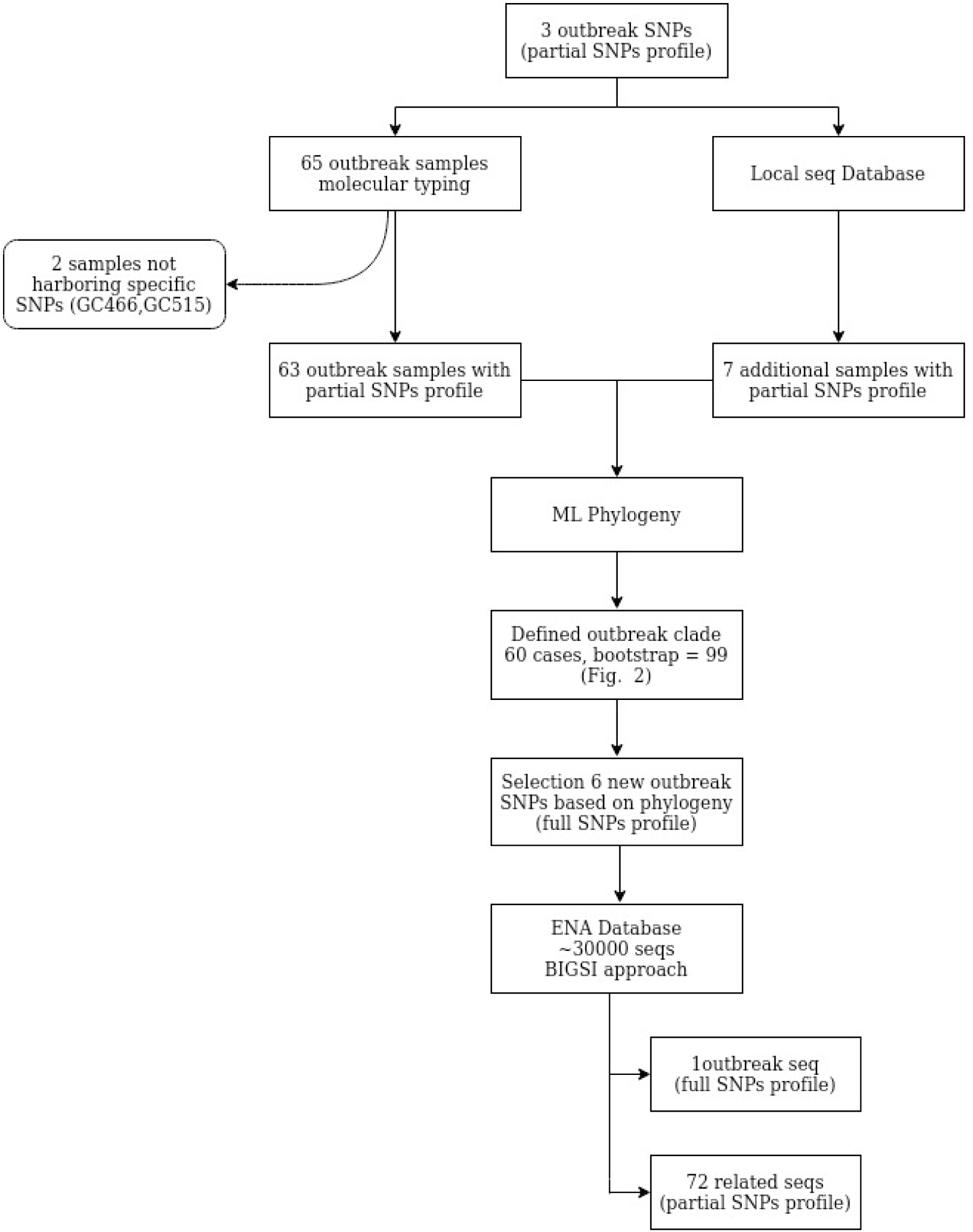
Workflow detailing the GC outbreak delineation procedure

First, all sequences were queried for the 3 SNP previously defined as GC outbreak markers (partial SNPs profile)[10]. We observed that 2 isolates (GC466, GC515), previously included in the outbreak based on molecular typing methods, did not actually harbor those marker SNPs, thus both were excluded from further analysis. In addition, we queried our collection of sequences and found 7 additional samples, 2 from Valencia Region and 5 from Liberia (Table S2). We then constructed a maximum likelihood (ML) tree with all the samples with the marker SNPs, and observed a differentiated and well-supported monophyletic clade (bootstrap = 99, Figure 2A) including the index case, most of the isolates previously assigned to outbreak and the Valencian cases (60 single TB cases, Table S1). The phylogeny also revealed that the SNPs initially considered as GC outbreak-specific, were not exclusive, since additional global strains also harbour them.

**Figure 2.**
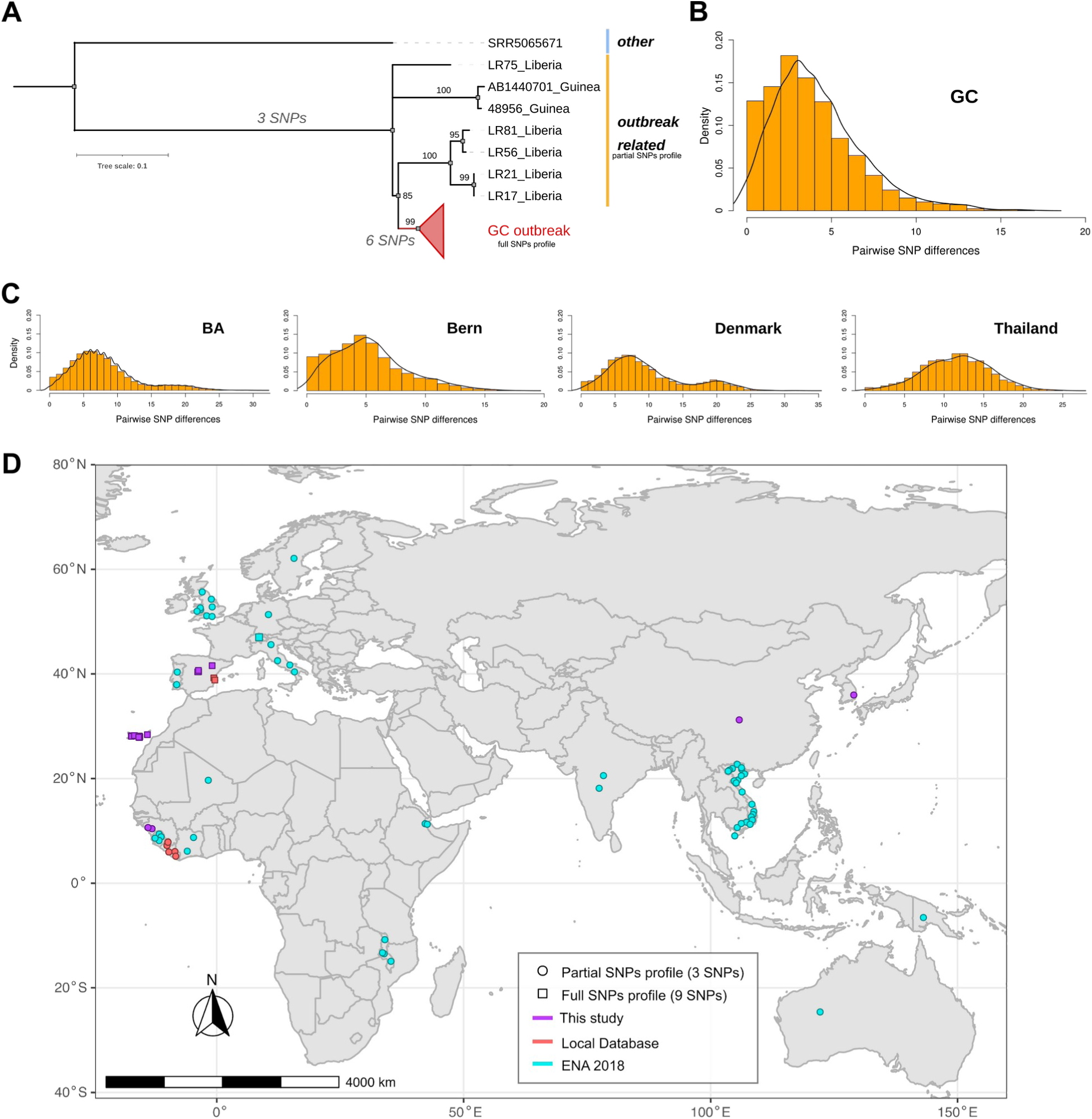
Genomic delineation of the outbreak. **A**. ML tree highlighting the phylogenetic circumscription of the outbreak and related strains identified with the partial SNPs profile. **B-C**. Density graphics of pairwise number of SNPs between samples of GC outbreak and Buenos Aires (BA) [5]; Bern [6]; Denmark [4]; Thailand [3]. **D**. Study of the success of GC outbreak outside the studied area, maps indicating the origin of the sequences identified by the different sources and their particular SNPs profiles; this study includes all samples sequenced as considered part of the outbreak by different molecular typing methods.

Pairwise distance was also estimated considering the phylogenetic outbreak definition based on WGS data (Table 1), in order to evaluate if distances were in agreement with the accepted recent transmission thresholds of 0-10 SNPs. Mean and median distance within-outbreak were 4.3 and 4 SNPs (range 0-17) respectively, while distance between outbreak and non-outbreak samples were 33 and 30 SNPs (range 18-63).

**Table 1.** Pairwise distance comparison among different outbreaks. Minimum (Min), maximum (Max), mean, median, 1^st^ (Q1) and 2^nd^ (Q2) quartile are provided

| Outbreak | SNP pairwise distance |  |  |  |  |  |
| --- | --- | --- | --- | --- | --- | --- |
|  | Min | Max | Q1 | Median | Mean | Q2 |
| GC | 0 | 17 | 2 | 4 | 4.3 | 6 |
| BA | 0 | 31 | 5 | 7 | 8.2 | 10 |
| Bern | 0 | 18 | 3 | 5 | 5.4 | 7 |
| Denmark | 0 | 33 | 6 | 8 | 9.9 | 13 |
| Thailand | 0 | 25 | 9 | 12 | 11.8 | 15 |

When compared to other known outbreak strains, the GC one displayed the lowest within-outbreak mean and median pairwise distance, though similar to the Bern outbreak (Table 1, Figure 2B-C). GC and Bern outbreaks exhibited a unimodal right-skewed pairwise genetic distance distribution, pointing out that most of the samples have low pairwise SNP distance among them. On the contrary, Denmark and Buenos Aires (BA) had a slightly bimodal distribution, suggesting the existence of more than one distinctive clade within both outbreaks, as proposed elsewhere [4]. Thailand outbreak displayed a unimodal normal distribution indicating a high pairwise SNP dispersion.

Thus, distance values obtained for GC also support the phylogenetic circumscription of the outbreak. With this delimitation, two additional samples from Madrid (AB1440701, 48956), belonging to recent Guinean immigrants, and previously identified as infected by the GC1237 strain [10], were excluded (Table S1).

### Evaluation of success outside the outbreak GC setting

In order to know if the GC strain can be found outside the Canary Islands, and if so if it has a similar epidemiological success, we designed an approach to evaluate all the MTB sequences available in public databases. This gives us the opportunity to study the spread of the outbreak, not only to the study area, but inspect its broader extension. First, the phylogenetic circumscription allows us to identify six additional markers defining the outbreak clade, most of which were missense mutations located in genes involved in metabolic processes and respiration, with effects likely unrelated with virulence or transmission (Table S3, FIgure 2A). Thus, the full SNPs profile (9 SNPs) was used to query every *M. tuberculosis* sample deposited in ENA by July 2018, using the BIGSI index [25] (see methods). We identified 1 sequence meeting the full SNPs profile, from Switzerland, thus being part of the outbreak, and 72 harboring the partial SNPs profile, thus being related to the outbreak (Table S2, Figure 2D). We consider these sequences in further analysis, as they can shed light on the remote origin of the outbreak strain.

### GC outbreak topology

To study the epidemiological characteristics of the GC strain in the Canary Island and abroad, we constructed a median joining network with all samples, including the three additional samples from the likely index case (Table S1, Figure 3). The genomic network displays a star-like structure. The likely index case is located in the center (including the oldest sample and two additional samples collected in 1994, 1995) along with 9 other isolates; most of them from 1993, and all collected from Gran Canaria. In depth investigation of central node (node A) revealed one heterogenous SNP (hSNP), at genomic position 3190007, for which the first sample of the likely index case (GC077) harbor both the reference (T: 82.7%) allele, shared by the rest of samples of the outbreak, and the alternative allele (C: 17.3%), which is fixed in sample GC092 from 1994 (Figure 3). The other hSNPs identified were uninformative since only one allele was fixed, either reference or alternative.

**Figure 3.**
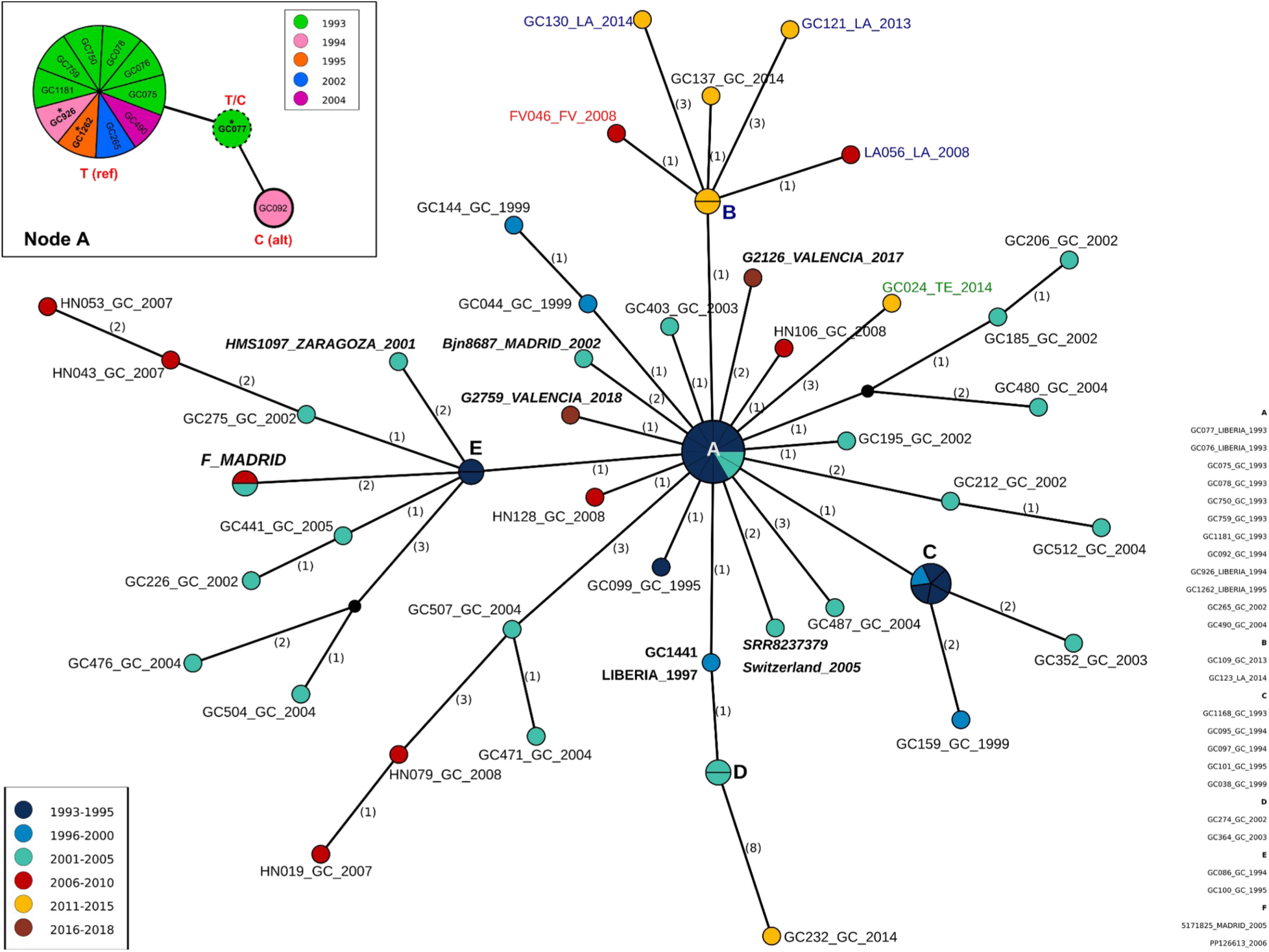
Median joining network analysis. Numbers in parentheses indicate the number of SNPs between nodes. Node size indicates the number of samples with the same genome, node color denotes sampling time; name color indicates different islands (blue: Lanzarote; red: Fuerteventura; green: Tenerife; black: Gran Canaria). Samples from the continent are indicated in italic and bold letters. GC1441 is an additional sample from the index case. Node A resolution with hSNPs pos: 3910007 is detailed; reference (ref) and alternative (alt) alleles distribution among samples is indicated. Names with asterisks indicate samples of the index case.

Connected with node A, three additional smaller star-like structures were observed, corresponding to nodes B, C and E (Figure 3), and resembling secondary outbreaks. Node B represents the later spreading (around 2008) from Gran Canaria to the other islands in the archipelago; with an additional independent introduction in Tenerife. Node C, the smallest secondary outbreak, also occurred in Gran Canaria. Notably, the biggest secondary outbreak, node E, started early and includes samples from the peninsula (Madrid and Zaragoza). Furthermore, the network reveals that the index case initiated a new transmission chain in 1997 (isolate GC1441), also in Gran Canaria. Surprisingly, the two samples from Valencia were directly linked to the origin of the outbreak, without connection between them. Epidemiological data agree with the absence of link between both cases, but there is also no evidence of trip to Canary Islands for either patient, suggesting an additional missing case (or cases) linking them. For the three isolates collected in Madrid, there is some link with Gran Canaria; case Bjn8687 stayed there in prison two years before diagnosis and the others (PP126613, 5171825) also visited the island at some point before diagnosis (Table S1). For the isolate identified from Switzerland, no epidemiological information was obtained. Overall, the extent of secondary transmissions after exportation events seems very limited, since no additional sequences from other places were retrieved from ENA.

### Phylodynamics analysis

We reasoned that if the GC strain had any transmission advantage this could be reflected in its natural history. Since we found evidence of temporal structure in our dataset (see Methods), it is suitable for applying a Birth-death skyline (BDSKY) model to estimate the epidemiological parameters of the GC outbreak (Supplementary Notes).

The becoming uninfectious or recovery rate (δ) resulted in a median value of 0.49 [0.28 - 0.75, 95% HPD] suggesting an infection period of 2 years [1.3 - 3.6 y] in agreement with the global estimation of 1-3 years previously proposed [35]. The effective reproduction number varies through the period of the outbreak, displaying a particular profile with peaks of expansion (R_e_>1) and reduction (R_e_<1). The peaks were observed approximately every ten years, and have an extension of three years (Figure 4A). Peaks matched with the secondary outbreak nodes observed in the network, and the periods of outbreak reduction coincide with chains instead of star-like transmission patterns (Figure 3). As observed in the histogram, peaks do not reflect the sampling effort, since periods 2005-2010 and 2010-2015 display a similar number of samples (Figure 4B). While in the first period isolates appeared in terminal nodes (Figure 3), in the second the cases are part of a secondary outbreak (Figure 3, node B). On average, the outbreak is decreasing, since the last period showed a R_e_ close to 1. Besides its particular profile, R_e_ never exceeds a value of 12 [6, 0.01 - 11 95% HPD], the maximum number of secondary cases caused by an infected person per year estimated for TB [35]. Overall, there is no indication from phylodynamics analysis that the GC strain has a transmission advantage, at least in terms of secondary cases generated.

**Figure 4.**
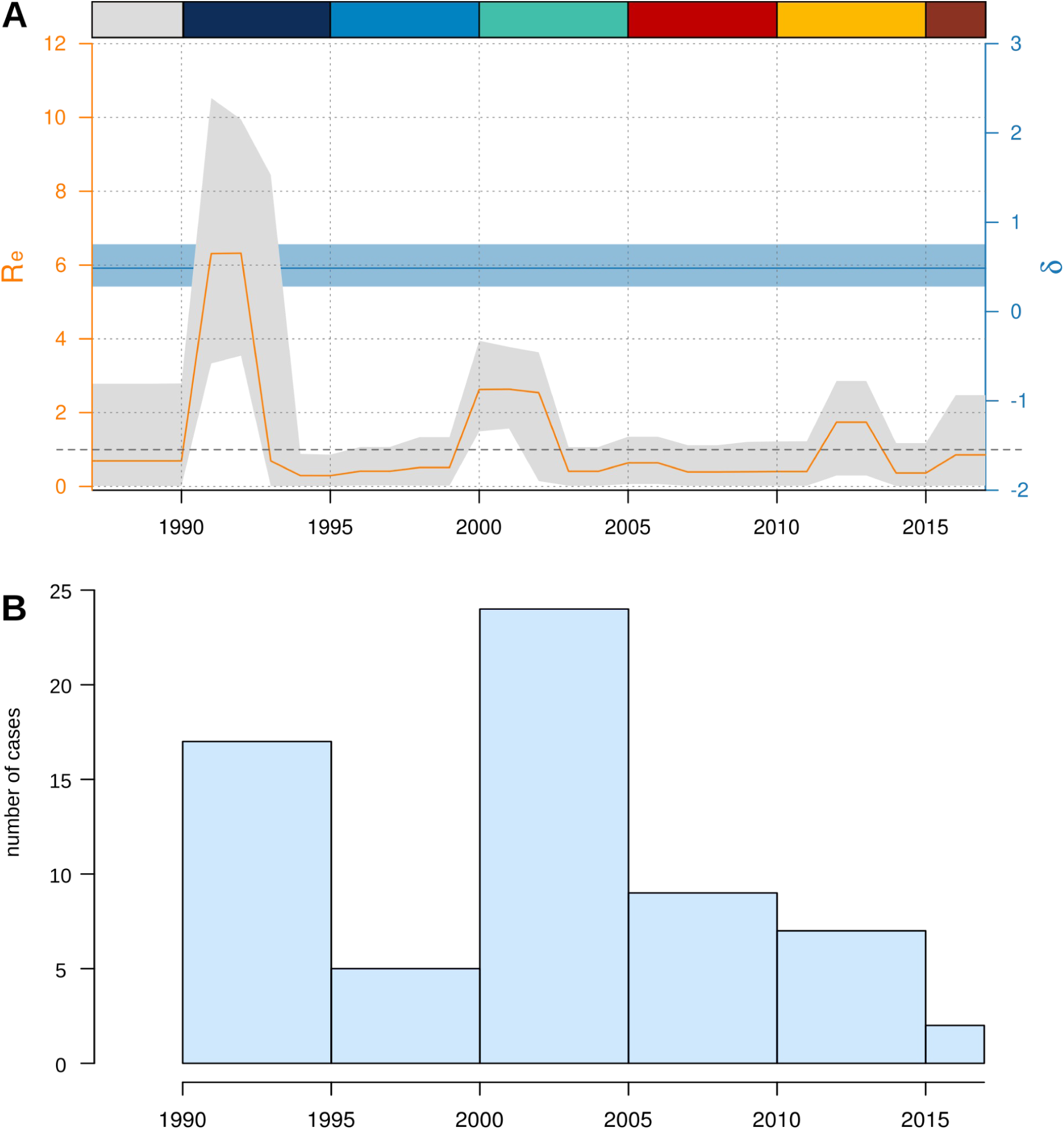
Phylodynamics analysis of GC outbreak. **A**. Birth-death serial skyline results showing reproductive number (R_e_) variation across outbreak period and recovery rate (δ). Rectangles in the top indicate periods of time as colored in Figure 3. **B**. Histogram of number of cases sequenced in the different time periods.

On the contrary, if ecological drivers like host or social determinants were behind the initial success of the GC strain, it will be mostly expected among the vulnerable population. In this sense, from all 61 patients belonging to the GC outbreak, 64% presented at least one risk factor including; PDU (Parenteral Drug Users), NPDU (Non-Parenteral Drug Users), imprisonment, HIV infection, alcohol abuse, indigence and no TB treatment adherence (Table S1). All of them are known risk factors for TB infection in low burden countries. Furthermore, considering only patients with available information, the value increases to 76%.

### Tracking the GC outbreak strain over centuries

The GC outbreak strain belongs to the MTBC lineage 2. A global ML phylogeny was constructed including 740 L2 global strains (MSA with 43401 concatenated SNPs) from different studies (Table S4). Placed in the context of L2 diversity and considering the classification previously proposed [38,39], the GC outbreak strain belongs to L2.2.3 (Figure 5A). The closest clade is mainly composed of African strains (Figure 5B, orange clade) followed by a mixed African-Vietnamese clade (Figure 5B, yellow clade) and more distantly related to China (Figure 5B). The emergence of the outbreak was estimated around 1984 AD [1973-1993 AD, 95% HPD] in agreement with the arrival time of the likely index case to the Archipelago (Figure S1). The MRCA of the African clade was estimated at 1930 AD [1913-1947 AD, 95% HPD] and the MRCA of the African-Vietnamese within the middle of 19th century (Figure 5B). Origin of L2.2 was estimated between 559-819 AD in agreement with previous estimates using the same approach [40]. Phylogeographic and dispersal reconstruction of the outbreak ancestors, placed its oldest ancestor in China around the middle 1300 AD; from there it spread to Vietnam between 1500-1700 AD. From Vietnam it dispersed to Africa between 1930-1940 AD, then to Canary Islands between 1960-1993 AD to later reach Europe between 1993-2002 (Figure 6, File S1).

**Figure 5.**
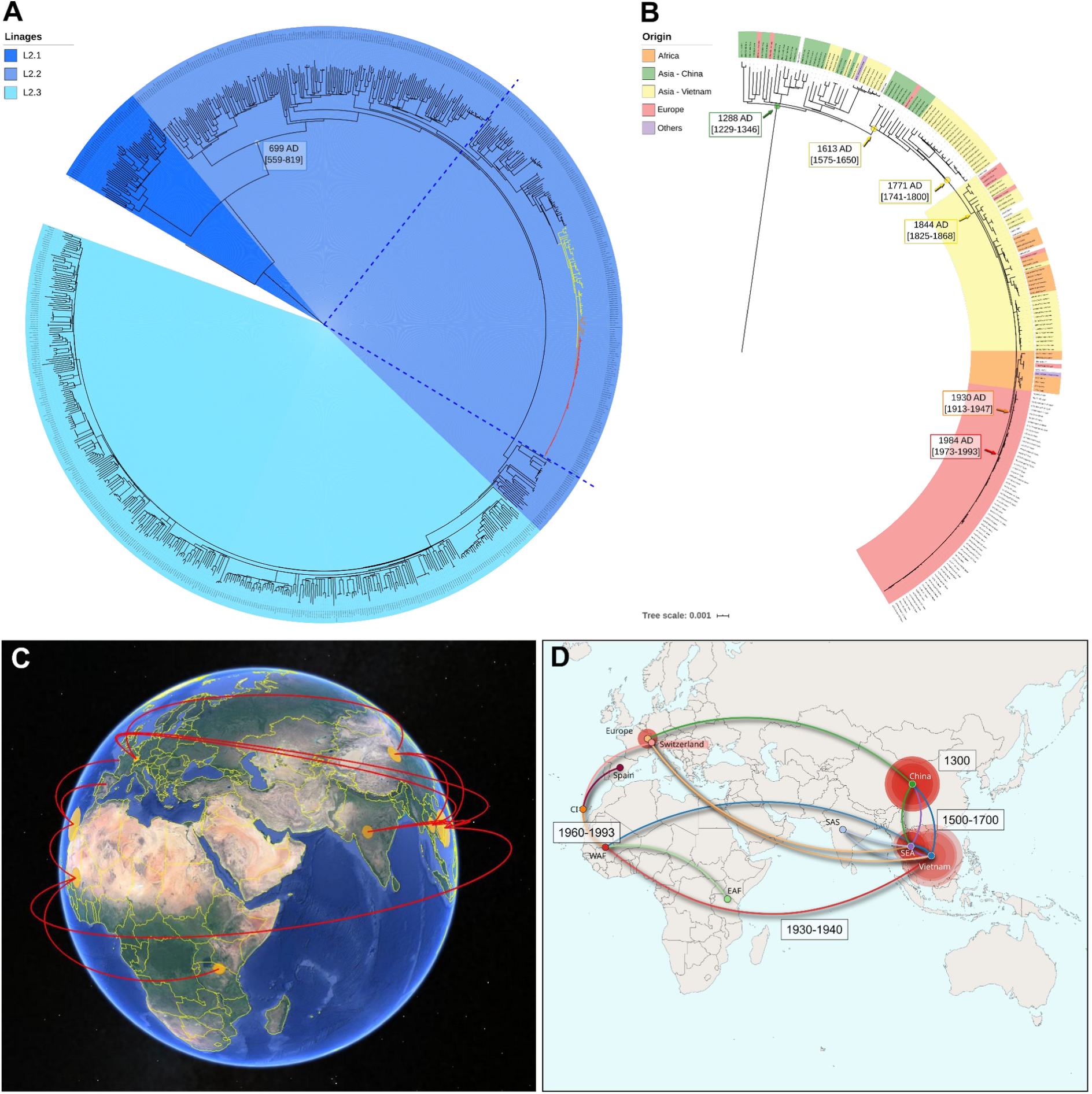
Dating, phylogeographic and dispersal analysis. **A**. ML global phylogeny of L2 highlighting the sublineages. Time of the most recent common ancestor (tMRCA) of L2.2. is indicated. Colored branches correspond to GC-outbreak (red), African related clade (orange) and African-Vietnamese clade (yellow). **B**. tMRCAs and origin of GC outbreak, closest nodes and oldest Chinese ancestor. Colors indicate origin of either samples (labels) and the ancestor of different clades. **C**. Results of the phylogeographic analysis in Google earth view. **D**. Routes of dispersion and time, lines are colored following the destination of the migration, and times from the oldest origin in China, to Vietnam and Canary Islands are denoted.

## Discussion

*Mycobacterium tuberculosis* is an obligate human pathogen and its success in the population deeply depends on human movement. By combining WGS and phylodynamics analyses, here we describe the dynamics of a 25 years outbreak in detail. After the index case arrival to Gran Canaria, the strain spread quickly, since many cases -with identical sequences- were observed in 1993. Two close secondary outbreaks appeared soon, one connected with the samples in the archipelago, and the other extended in the same island, probably as an early independent transmission chain. Lately, a differentiated secondary outbreak spread to the rest of the islands, reaching high representativeness [12]. By evaluating serial samples from different years, we could identify secondary outbreaks associated with the likely index case, probably due to poor treatment adherence. The deep analysis of hSNPs resolved the relations among samples in the central core of the outbreak, adding molecular evidence to support GC077 as the index case (first sample). A similar approach has been described in Lee et al. [22] showing the potential of using hSNP for further genetic discrimination within outbreaks. This approach also provided evidence that most of the observed diversity within the outbreak was already present in the index case samples. Furthermore, this approach allowed us to circumscribe the outbreak, discarding distantly related isolates linked by molecular methods that only query a restricted portion of the genome.

With the added value of querying a large collection of WGS [13,25], our comprehensive approach allowed us to link the GC strain to cases in continental Europe and thus determine the extent of an outbreak beyond the geographic limits where it was studied. Thus, while the GC outbreak strain had a very high local success, reaching a frequency of 27% of all isolates in Gran Canaria [12], it had limited expansion outside the archipelago; with multiple introductions in the continent not resulting in secondary cases [10].

By combining the finding of a wide SNP profile and by querying a large dataset, we were able to shed light on the remote origin of the outbreak strain. Our analysis places the ancestors of the GC strain hundreds of years ago, in China around 1300 AD. Thanks to the vast amount of MTBC genomic data available we have traced its initial movements from China to Vietnam, later to Liberia and finally to Gran Canaria by the end of the 20^th^ century. Once in Gran Canaria the strain generated a large number of secondary cases there, but was rarely seen outside the archipelago. Thus, here we were able to document how past epidemiological events impact current TB epidemics.

In addition to inferring its remote origin, we had the unique opportunity to study its proximal origin using a phylodynamic approach. Proximal origins of large TB outbreaks are rarely identified by epidemiological information, mainly because of the difficulty in identifying the index patient in an area with many local cases and, in the case of tuberculosis, because of latency periods. Since the GC outbreak was undoubtedly linked to the migration of an infected person from Liberia to Gran Canaria in 1993 [9], this represents a unique opportunity to validate the commonly applied Bayesian approach for tuberculosis. Our results (1984 AD [1973-1993 AD, 95% HPD]) largely agreed with the available epidemiological information.

The main limitations of our study is the low proportion of outbreak cases sequenced from Canary Islands, it accounts for approximately 10% of total cases, however we show that the dataset has enough phylogenetic signal for the analysis presented. In accordance, our phylodynamics results do not correlate to sampling effort (Figure 4). Spain also lacks a systemic WGS program, however in our study we use systematic typing data from three of the largest regions in Spain: Madrid, Aragón and Valencia Region to confirm that the strain had no success beyond the archipelago. Finally, by querying the whole ENA database we could retrieve additional cases sequenced elsewhere and could evaluate the expansion of the outbreak.

A common theme when it comes to MTBC strains is whether some are more transmissible than others or if its success is driven by ecological factors, or both. Outbreak strains are good candidates to look for intrinsic transmissibility as they are responsible for a large number of cases with sustained transmission over decades. For the GC strain, we estimate an infection period close to two years and less than 12 secondary cases per infected individual, same values proposed for TB in general [35], suggesting that there is no transmissibility advantage of the GC strain, at least in terms of shorter latency periods or higher contagion rate. In addition, it was demonstrated that GC strain did not display higher virulence, nor accumulated mutations in sequential samples or acquired resistance mutations [11]. Similarly, our extensive query of outbreak sequences in public repositories did not identify that this strain is responsible for outbreaks outside the Canary Islands. For example, an in-depth genotyping effort did not identify secondary cases associated with a case in Madrid that had prolonged disease [11] or in Aragón where systematic genotyping is applied to all TB positive cases. In addition, mutations associated with the GC strain are unlikely to have a role in transmission. For other outbreak strains, notably the Danish C2 [41] and the Toronto [42] strains, mutations with a functional role have been proposed but no experimental evidence is available. On the contrary, all those outbreak strains share the fact that they thrive in populations with significant risk factors, particularly during the ‘80s and ‘90s of the past century. Overall, data suggests that the success of the long-lived GC outbreak strain, and probably others, is related to ecological factors associated with founder effects linked to the host and social determinants of the disease rather than an intrinsic transmissibility difference.

## Data Availability

Availability of different type of data is described in detail in the mansucscript

## Acknowledgements

This project has been funded by the European Research Council (101001038-TB-RECONNECT), the Ministerio de Economía, Industria y Competitividad (PID2019-104477RB-I00) and European Commission –NextGenerationEU (Regulation EU 2020/2094), through CSIC’s Global Health Platform (PTI Salud Global) to IC. This project has been funded by the Instituto de Salud Carlos III (FIS18/0336), Gobierno de Aragón/Fondo Social Europeo “Construyendo Europa desde Aragón” to SS.

## Contributors

M.G.L and I.C conceived the study; M.I.C.H, F.C, R.C, L.S, B.P, L.P.L, D.G.V, S.S, J.C., E.L. collected the samples, obtained and curated the epidemiological data; M.T.P. processed and sequenced the samples; P.W., Z.I performed BIGSI analysis; M.M.M. performed the scripts for DST analysis; M.G.L performed all analyses; M.G.L and I.C wrote the first draft of the manuscript, with revisions from all co-authors.

