## Supplemental information for "Deciphering the tangible spatio-temporal spread of a 25 years tuberculosis outbreak boosted by social determinants"

*Temporal signal analysis*

Before applying the phylodynamics approach to study the evolution of the outbreak, we evaluated the clock signal of our dataset and compared it with other TB outbreaks, previously published. A moderate but significant clocklike structure was obtained with TempEst analysis ($R^2$=0.32, p-value<0.0001), stronger in comparison with outbreaks from Denmark ($R^2$=0.23, p-value<0.0001(Folkvardsen et al. 2017)); Switzerland ($R^2$=0.05, p-value<0.0001(Stucki et al. 2015)); Thailand ($R^2$=0.14, p-value = 0.011(Coscolla et al. 2015)) and Argentina ($R^2$=0.08, p-value<0.0001(Eldholm et al. 2015)) (Figure S2). Date randomization test (DRT) was also performed, it involves randomly reassigning sampling times of sequences. It was repeated 100 times, and the same BDSKY analysis described in methods was performed for each replicate. The simple DTR was passed since the median value of clock rate for the observed data does not overlap with the 95% HPD (highest posterior density interval) from the randomized test (Figure S3). There is only one randomized replicate overlapping with observed data, but it is the result of dates similarity among samples, this replicate is very similar to the observed data. The results provide evidence for temporal structure in our dataset. (Folkvardsen et al. 2017)

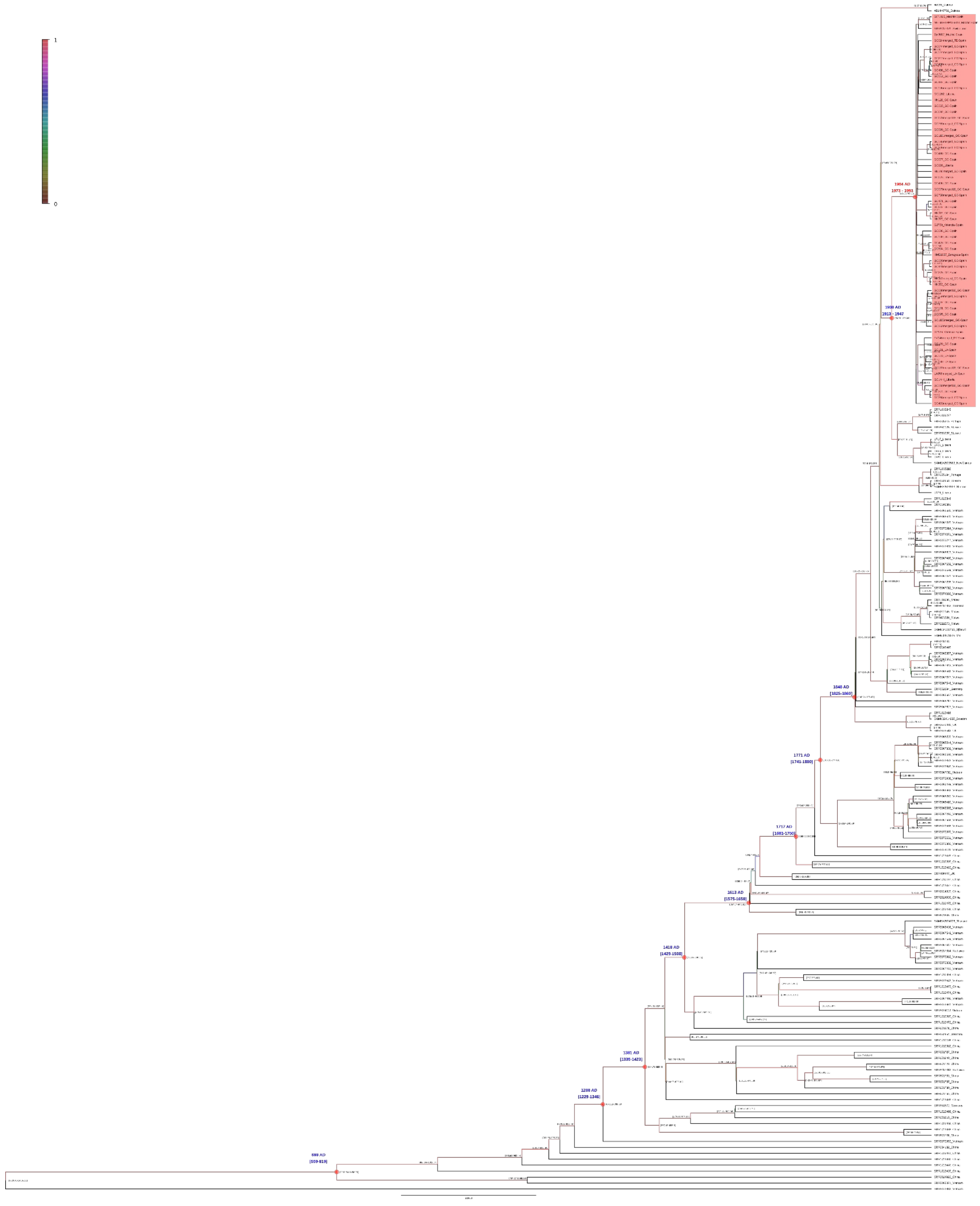
1
0
1984 AD
1973 - 1993
1913 - 1947
1848 AD
[1825-1860]
1771 AD
[1741-1800]
1717 AD
[1681-1750]
1613 AD
[1575-1650]
[1429-1508]
1381 AD
[1336-1423]
1288 AD
[1229-1346]
[559-819]

**Figure S1.** Maximum clade credibility tree obtained with Beast. 95% Highest probability density intervals (95% HPD) are indicated for all nodes. Median time and 95%HPD of the most recent common ancestor of the GC outbreak (in red) and related nodes (in blue), are indicated. Color branch denotes posterior values as indicated in the color scale.

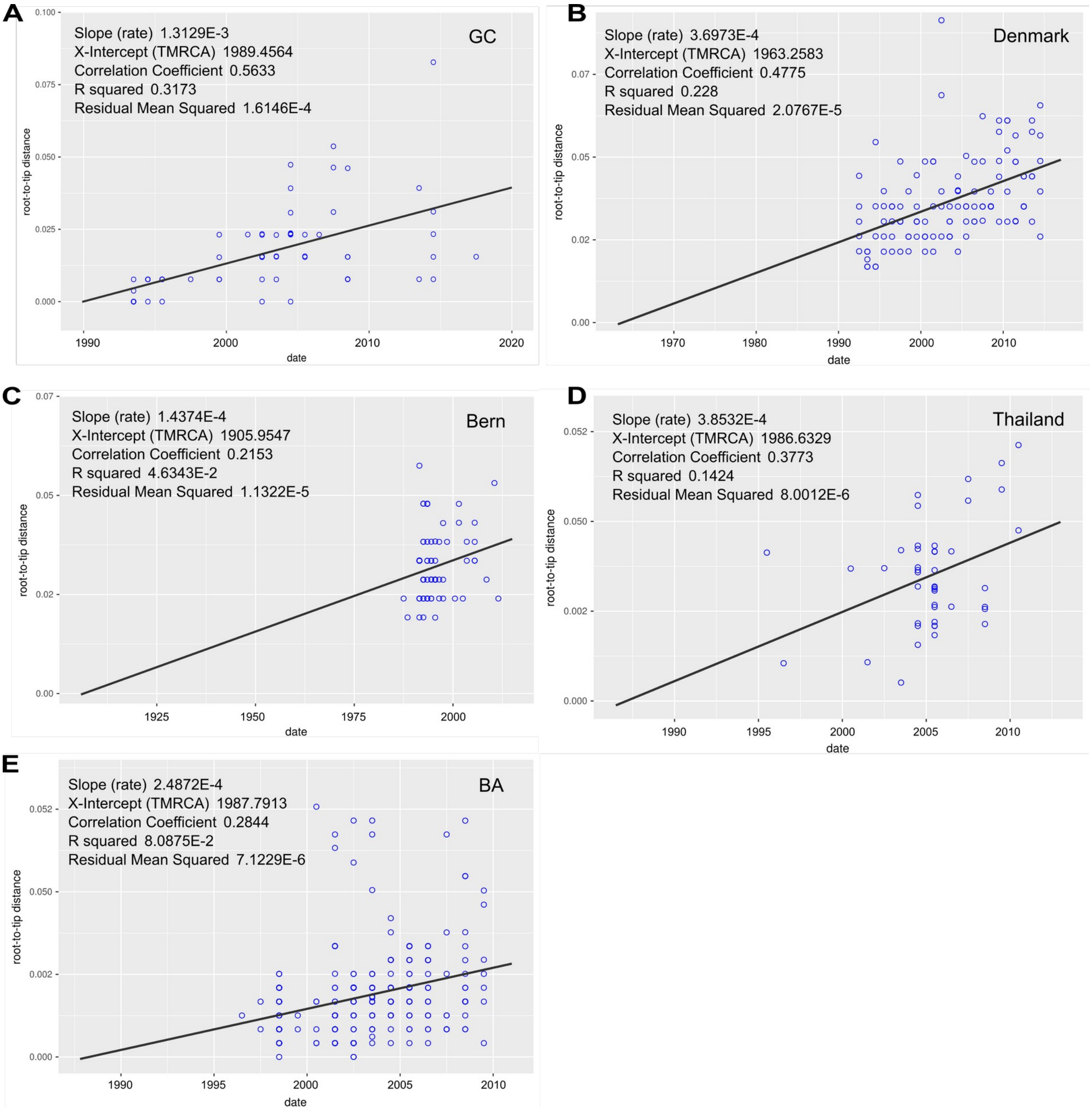


**Figure S2.** Root to tip analyses obtained with Tempest for GC (A), Denmark (B), Bern (C), Thailand (D) and Buenos Aires (E) outbreaks. Clock rate (slope), time of the most recent common ancestor (TMRCA) and Rsquared are indicated in each plot.

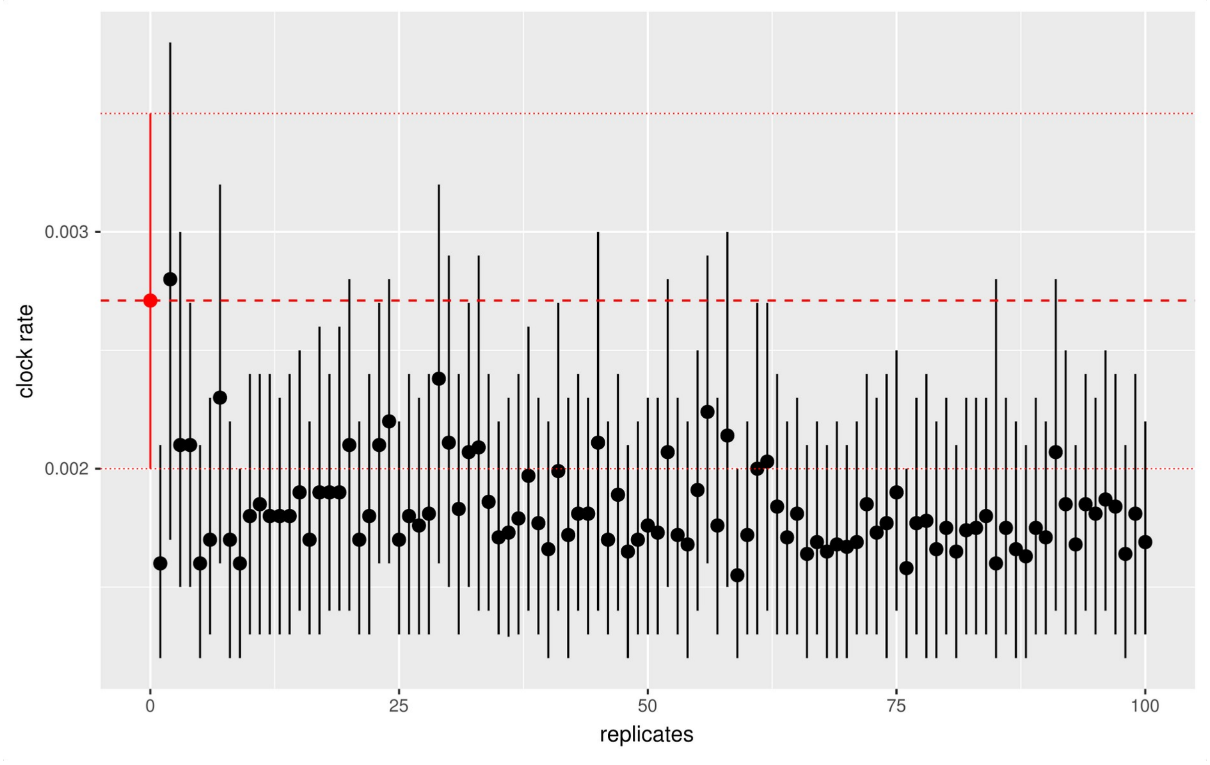


**Figure S3.** Date randomization test (DRT) performed for GC outbreak. Clock rate with 95% Highest posterior density interval is plotted for each of the 100 replicates with dates randomized. In read the values obtained with the observed data.
